# Multiple-time measurements of multidimensional psychiatric states from immediately before the COVID-19 pandemic to one year later: A longitudinal online survey of the Japanese population

**DOI:** 10.1101/2021.08.01.21261309

**Authors:** Taiki Oka, Takatomi Kubo, Nao Kobayashi, Fumiya Nakai, Yuka Miyake, Toshitaka Hamamura, Masaru Honjo, Hiroyuki Toda, Shuken Boku, Tetsufumi Kanazawa, Masanori Nagamine, Aurelio Cortese, Minoru Takebayashi, Mitsuo Kawato, Toshinori Chiba

## Abstract

The coronavirus disease 2019 (COVID-19) pandemic has profoundly affected the mental health of both infected and uninfected people. Although most psychiatric disorders have highly overlapping genetic and pathogenic backgrounds, most studies investigating the impact of the pandemic have examined only single psychiatric disorders. It is necessary to examine longitudinal trajectories of factors that modulate psychiatric states across multiple dimensions. 2274 Japanese citizens participated in online surveys presented in December 2019 (before the pandemic), August 2020, Dec 2020, and April 2021. These surveys included nine questionnaires on psychiatric symptoms, such as depression and anxiety. Multi-dimensional psychiatric time series data were then decomposed into four principal components. We used generalized linear models to identify modulating factors for effects of the pandemic on these components. The four principal components can be interpreted as general psychiatric burden, social withdrawal, alcohol-related problems, and depression/anxiety. Principal components associated with general psychiatric burden and depression/anxiety peaked during the initial phase of the pandemic. They were further exacerbated by the economic burden of the pandemic. In contrast, principal components associated with social withdrawal showed a delayed peak, with human relationships as an important risk modulating factor. In addition, being elderly and female were risk factors shared across all components. Our results show that COVID-19 has imposed a large and varied burden on the Japanese population since the commencement of the pandemic. Although components related to the general psychiatric burden remained elevated, peak intensities differed between components related to depression/anxiety and those related to social anxiety. These results underline the importance of using flexible monitoring and mitigation strategies for mental problems, according to the phase of the pandemic.

## INTRODUCTION

The coronavirus disease 2019 (COVID-19) pandemic has affected all aspects of society globally [1, 2]. Given its profound impact on the mental health of both infected and uninfected persons, there is a greater need for mental health science [3–6]. Indeed, various psychiatric states, such as depression [7], general anxiety [8], panic disorder [8], social anxiety disorder [9], alcohol [10], internet-related problems [11], adult attentiondeficit/hyperactivity disorder (ADHD) [12], and autism spectrum disorder (ASD) [13] have been exacerbated during COVID-19. To prevent further deterioration and persistence after the pandemic subsides, a comprehensive understanding of longitudinal trajectories of psychiatric states during this pandemic is required.

To reach a full grasp of pandemic effects on mental health, it is necessary to evaluate data that include prepandemic data as a baseline, as well as multiple time-points during the pandemic, and multiple psychiatric states. Baseline control data are essential to directly assess any pandemic effects. Most evidence to date is based on crosssectional samples and very few studies have included data from immediately before the pandemic [10, 14–17]. Second, multiple time points during the COVID-19 pandemic should be included. This is because some psychiatric symptoms, such as depression and anxiety, react to the initial stages of natural disasters, including the pandemic [7, 18–20], while others, such as suicide, show delayed reactions [21–23]. Indeed, both patterns have been observed during this pandemic. Third, multiple psychiatric states should be addressed simultaneously in the same population, since psychiatric disorders have highly overlapping genetic backgrounds and pathogenesis [24, 25], and a shift from categorical to dimensional classification of psychiatric disorders has long been advocated [26, 27]. Generally speaking, psychiatric disorders can be decomposed into three factors that interact strongly; 1) anxiety and depression (internalizing), 2) substance dependence, antisocial personality disorder, and conduct disorder (externalizing), 3) bipolar disorder and schizophrenia (psychotic experience) [28–30]. Furthermore, it has also been proposed that these three latent traits represent varied manifestations of a single, general, psychopathological dimension, called the “*p* factor” [24, 31, 32]. Is it possible then, to decompose psychiatric disorders into several (or even single) factors or components, depending on how they have been affected by the COVID-19 pandemic?

By analyzing data before and during the pandemic, and by gathering data about nine psychiatric states, we identified four underlying principal components (PCs). Of these, three PCs showed distinct exacerbation trajectories, with different risks and protective factors. Results of this study may help to optimize strategies to improve mental well-being in at-risk populations.

## METHODS

### Procedures and outcomes

We conducted a repeated online survey with the help of Macromill Inc. (Japan) (See Supplementary Methods for details). The original survey was conducted in December 2019. Since the first COVID-19 case was not identified in Japan until January 2020, data from this survey are considered baseline data (T0). In response to the COVID-19 pandemic, individuals from this survey were invited to participate in follow-up surveys that took place in August 2020 (T1), December 2020 (T2), and April 2021 (T3). In these follow-up surveys, several items were added regarding COVID-19. Multidimensional psychiatric data taken immediately before and during COVID-19 were collected from a total of 4680 participants. We excluded 326 individuals because of inconsistencies or contradictions in their answers (details in Supplementary Methods). An additional 419 individuals were excluded because they responded identically to all items, using only the maximum or minimum values in the questionnaires, including reverse questions. Among the remaining 3935 individuals, 1661 participated in only two or three surveys. In the end, 2274 individuals who responded to all four surveys comprised the main study population. We repeated the main analyses with data that included partial participants to rule out confounds due to dropout from the surveys (N = 3935; survey population). (Figure 1). The original and follow-up research designs were approved by the Ethics Committees of the Advanced Telecommunications Research Institute International (ATR) (approval No. 21-195 for the original study & 21-749 for the follow-up study). All participants read a full explanation of the study and gave informed consent before each survey.

**Figure 1.**
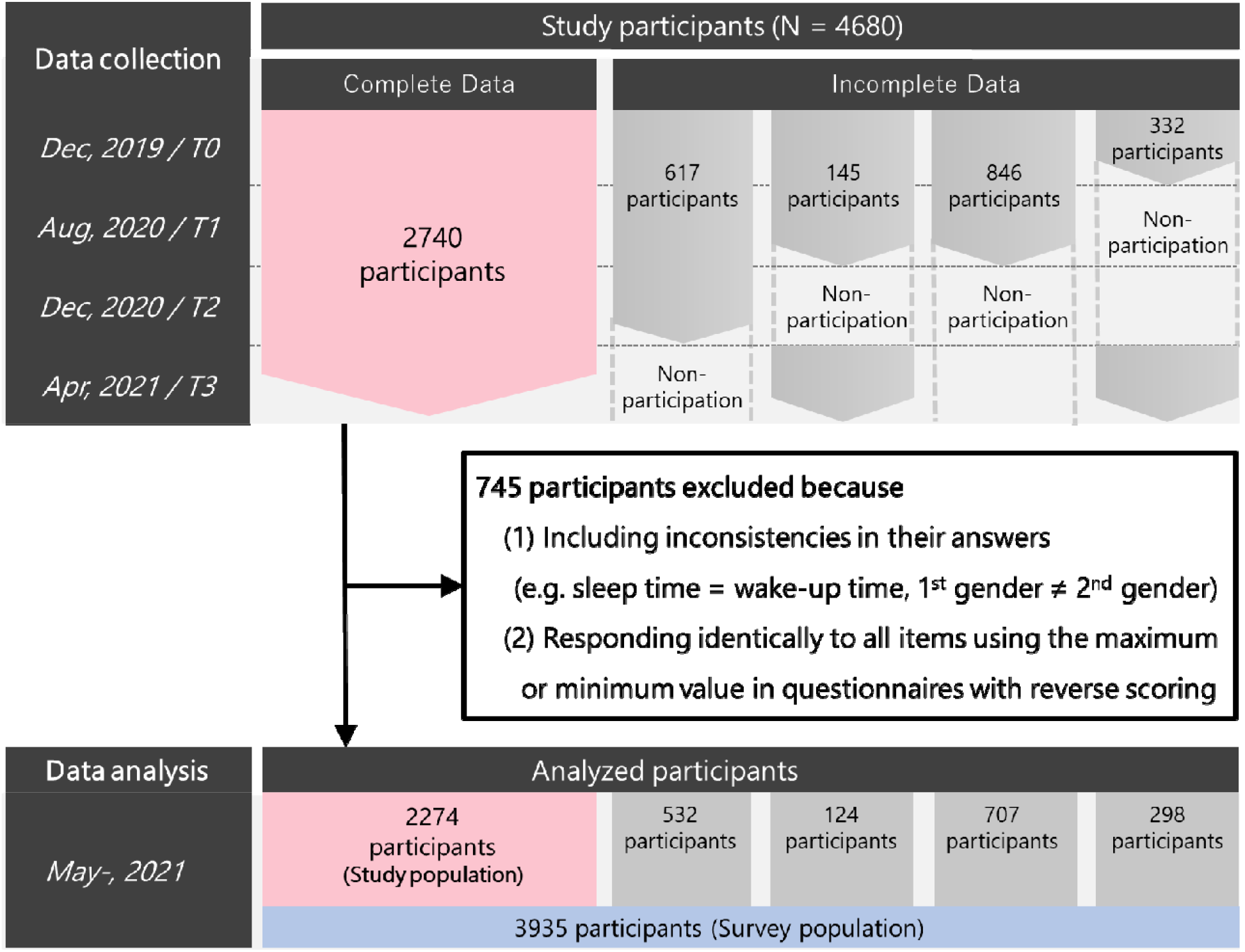
Chart showing selection into cohorts 4680 participants participated in the T0 survey immediately before and during COVID-19. 745 participants were excluded due to data problems and 3935 participants remained in the analysis (defined as “Survey population”). 2274 of those responded to all four surveys (defined as “Study population”). Of the 3935, 1661 participated in only two or three surveys (incomplete data) (532, T0-T2; 124, T0-T1, T3; 707, T0-T1; 298, T0 and T3).

Demographic variables were collected as follows: gender (women and men), age, job status (Self-employed, employed, unemployed and other), education history, household income per year (<4 million yen, 4-6 million yen, 6-8 million yen, 8-10 million yen, >10 million yen, and missing), the age of the youngest child in the household (none; 0–3; 4–6; 7–12;13–15; 16–18; ≥18), and the number of cohabitants (alone; 2–3; ≥4).

We assessed the psychiatric status of each respondent using validated questionnaires, evaluating them for major depressive disorder (CES-D) [33], obsessive-compulsive disorder (OCI) [34], internet-related problems (CIUS) [35], attention-deficit/hyperactivity disorder (ADHD) (ASRS) [36], autistic spectrum disorder (AQ) [37], social anxiety (LSAS-fear/avoid) [38], general anxiety (STAI-Y-state) [39], and alcohol-related problems (AUDIT) [40]. Details for each questionnaire are in the Supplementary Methods. COVID-19-related items were also measured in surveys during the pandemic (Supplementary Table 1 and Supplementary Methods for details about the survey).

### Statistical analysis

Each disorder score at T0 was normalized across subjects. Scores at different time points were standardized in addition to the mean and variance at T0. Next, normalized scores from each participant were concatenated temporally. Correlation among all disorder pairs of concatenated score vectors were calculated to check the covariance of psychiatric status. Then, to extract principal components of multidimensional psychiatric disorders, we performed a principal component analysis on the concatenated data. Using an orthogonal transformation, a set of correlated dimensions were converted into a set of uncorrelated components [41]. Hence, each principal component (PC) consisted of a set of correlated components of psychiatric disorders, i.e., that covaried over time during the pandemic. How much each disorder contributed to each PC was numerically indexed as the “loading” of that disorder. The top four PCs explained ∼60% of the variance in the data. The nine psychiatric scores of each participant at each time point were then converted into these four PCs, based on the loadings. Changes in scores of each psychiatric disorder and each PC across time-points were tested by one-way ANOVA and Tukey’s test. Multiple comparisons were adjusted using Bonferroni correction. The adjusted *p*□<□0.05 (i.e., unadjusted *p*□<□0.0056 in each psychiatric analysis, unadjusted *p*□<□0.0016 in each PC analysis) was considered the threshold for statistical significance.

To identify risks and protective factors against exacerbation of each PC, we performed generalized linear model regression. In each regression model, changes in each component from the baseline for each participant were used as dependent variables. Independent variables included demographics (gender, age, number of cohabitants, marital status, age of youngest child, income, and job type), COVID-19 related variables (income changes, number of received government compensation payments, and changes in daily communication frequency both online and face-to-face, see Supplementary Table 1) and all four PC scores at pre-pandemic (T0).

## RESULTS

We analyzed data from 2274 study participants who participated in all 4 surveys. Demographic data of participants are provided in Table 1. Characteristics of the survey population (N = 3935, which includes participants who did not complete all four surveys) are described in Supplementary Table 2.

**Table 1.** Characteristics of study population (N = 2274)

|  |  |  |  |  |  |  |  |  |
| --- | --- | --- | --- | --- | --- | --- | --- | --- |
| All | 2274 (100%) |  |  |  |  |  |  |  |
| Gender | Male | Female |  |  |  |  |  |  |
|  | 1225 (54%) | 1049 (46%) |  |  |  |  |  |  |
| Marital status | Not married | Married |  |  |  |  |  |  |
|  | 801 (35%) | 1473 (65%) |  |  |  |  |  |  |
| Age of youngest child in household | No children | 0~3 | 4~6 | 7~9 | 10~12 | 13-15 | 16-18 | 19~ |
|  | 1254 (55%) | 160 (7%) | 90 (4%) | 101 (4%) | 106 (5%) | 100 (4%) | 101 (4%) | 362 (16%) |
| Household income | Lowest | 2nd | 3rd | 4th | Highest | Missing |  |  |
|  | 567 (25%) | 495 (22%) | 409 (18%) | 222 (10%) | 382 (17%) | 199 (9%) |  |  |
| Job | No job | Self-employed | Employee | Other |  |  |  |  |
|  | 375 (16%) | 517 (23%) | 1177 (52%) | 205 (9%) |  |  |  |  |

Correlation matrices for changes in scores of psychiatric disorders over time are shown in Supplementary Figure 2. Half (18/36) of all combinations had relatively strong positive (>0.5) or negative (< -0.5) correlations.

### Impact of the pandemic on scores of each psychiatric disorder

During the observation period, we found abrupt, statistically significant exacerbation of general anxiety, avoidance aspects of social anxiety disorder, and internet-related problems (Supplementary Table 3). These disorders showed no remission during the study. A similar, but non-significant pattern was observed for autism spectrum disorder (ASD). The fearful aspect of social anxiety disorder, obsessive-compulsive disorder (OCD), and attention-deficit/hyperactivity disorder (ADHD) showed a statistically significant initial drop followed by a gradual increase. Major depressive disorder and alcohol-related problems showed an initial increase, followed by a gradual decrease (Figure 2A; Supplementary Figure 2).

**Figure 2.**
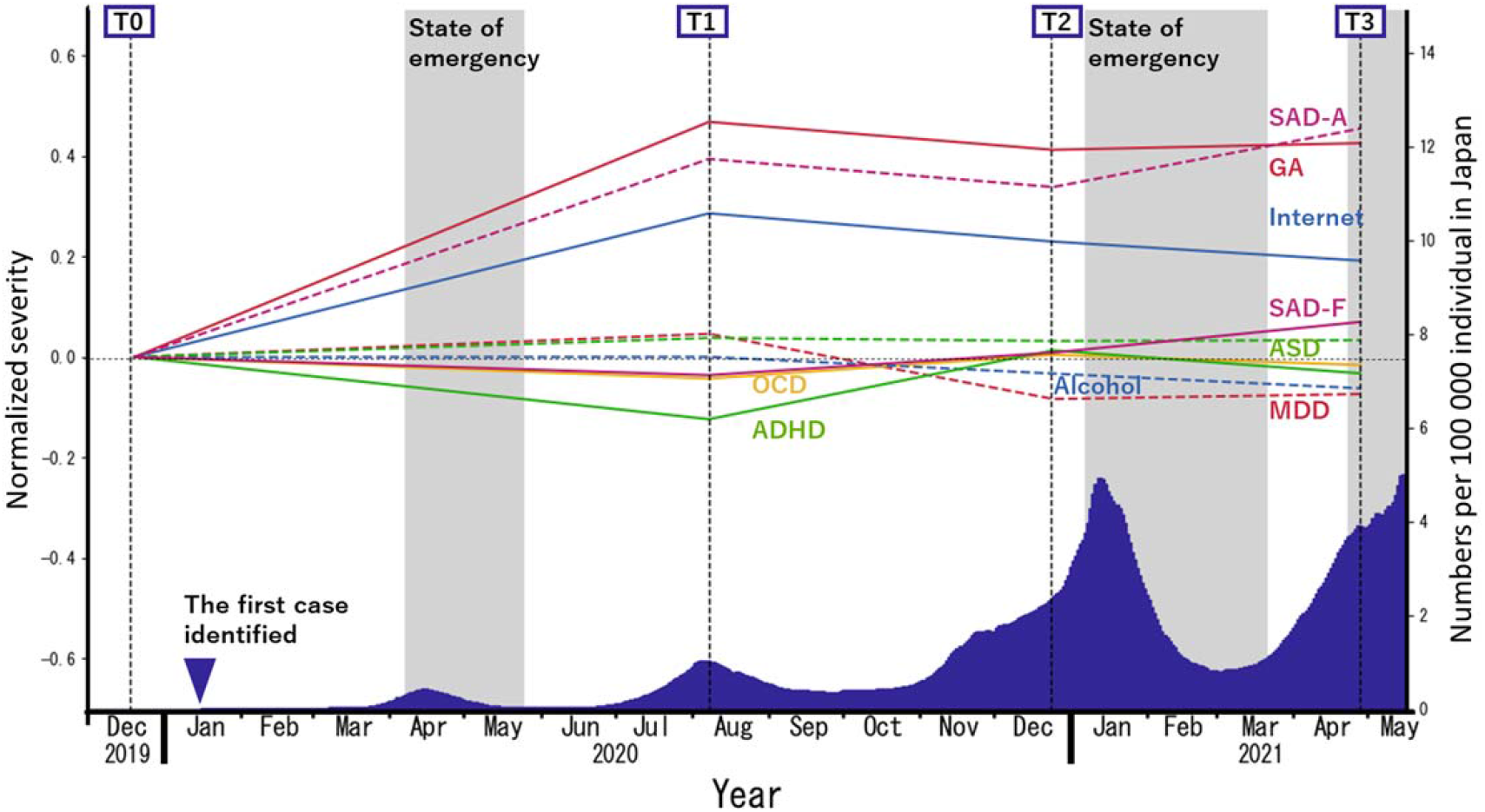
Trajectories of psychiatric scores in the study population (N = 2274) The blue area under the curve represents a 15-day moving average of daily new cases of COVID-19 per 100 000 Japanese residents. GA: general anxiety as measured by STAI-Y; SAD-A: avoidance aspects of social anxiety disorder as measured by LSAS-A; Internet: internet-related problems as measured by CIUS; ASD: autism as measured by AQ; SAD-F: fear aspects of social anxiety disorder as measured by LSAS-F; OCD: obsessive-compulsive disorder as measured by OCI; ADHD: attention deficit and hyperactivity disorders as measured by ASRS; Alcohol: alcohol-related problems as measured by AUDIT; MDD: major depressive disorder as measured by CES-D.

### Principal component analysis of multi-dimensional psychiatric states

Multi-dimensional psychiatric time-series data were decomposed into orthogonal principal components. The top 4 components explained approximately 60% of the variance (PC1: 24.1%, PC2: 14.3%, PC3: 11.0%, PC4: 10.6%, in total 59.9%), so we analyzed those further. Principal component 1 (PC1) included all psychiatric illnesses. Principal component 2 (PC2) was composed of social anxiety and internet-related problems. Principal component 3 (PC3) consisted mainly of alcohol-related problems. Principal component 4 (PC4) comprised depression, anxiety, and alcohol-related problems (Figure3 A). Characteristics of each PC loading are shown in Supplementary Table 4 for the study population and in Supplementary Table 5 for the survey population. All but PC3 worsened during the initial phase of the pandemic, followed by further exacerbation (PC2), sustained elevation (PC1), and partial remission (PC4) (Figure3 B). Despite showing slight remission in later phases (T2, T3), comparable to the initial phase (T1), PC4 remained higher than baseline (T0) throughout the pandemic. PC3 was excluded from further analysis to identify participant risk/protective factors because throughout the pandemic, it showed no significant changes relative to baseline (T0) (Supplementary Table 6; trajectories of each PC in survey population are shown in Supplementary Figure 3).

**Figure 3.**
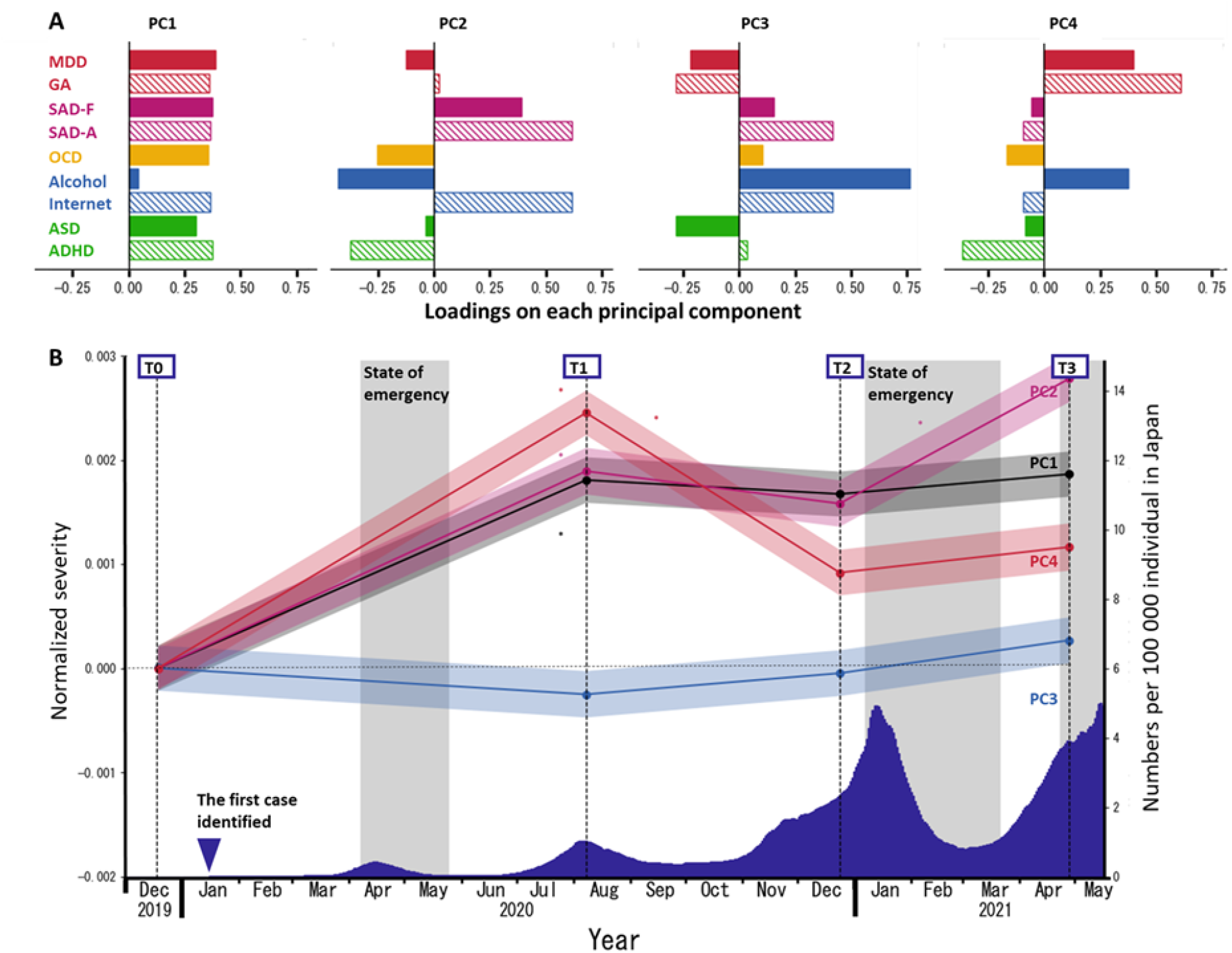
Trajectories of PC scores obtained from multidimensional psychiatric scores in the study population (N = 2724) **A**. Loadings of psychiatric disorder scores for each principal component. **B**. Trajectories of the average of each PC are shown. The blue area under the curve represents a 15-day moving average of daily new COVID-19 cases per 100 000 Japanese residents. Signs of PCs were arranged so that each maximum loading assumed a positive value. Asterisks indicate significant changes in PC score from the previous time point (p < .05, Bonferroni-corrected). All PC scores during the pandemic (T1, T2, and T3) except those of PC3 are significantly higher than the scores prepandemic (T0). MDD: major depressive disorder as measured by CES-D; GA: general anxiety as measured by STAI-Y state, SADF: fear aspects of social anxiety disorder as measured by LSAS-F; SAD-A: avoidance aspects of social anxiety disorder as measured by LSAS-A; OCD: obsessive-compulsive disorder as measured by OCI; Alcohol: alcohol-related problems as measured by AUDIT; Internet: internet-related problems as measured by CIUS; ASD: autism as measured by AQ; ADHD: attention deficit and hyperactivity disorders as measured by ASRS

### Regression analyses to identify risk and protective factors

Figure 4 shows the results of generalized linear models to identify factors that may exacerbate or alleviate effects of the pandemic on components of each PC. All results are reported with coefficients ± standard error (SE) and p-values (*p*). In PC1 and PC2, being female represented a significant risk of exacerbation compared to being male (*β* = 0.32 ± 0.05, *p* < 0.001; *β* = 0.38 ± 0.04, *p* < 0.001). Older age was also associated with exacerbation of PC2 (*β* = 0.07 ± 0.02, *p* = 0.010). In PC1 and PC4, the impact of the pandemic on decreased household income was a risk factor (*β* = 0.08 ± 0.02, *p* < 0.001; *β* = 0.06 ± 0.02, *p* < 0.007). On the other hand, in PC2, being self-employed, experiencing changes (both increase and decrease) in face-to-face communication-time with family, and decreased online communication time with family were protective factors (*β* = -0.16 ± 0.06, *p* = 0.017; *β* = -0.14 ± 0.05, *p* = 0.007; *β* = -0.12 ± 0.05, *p* = 0.034; *β* = -0.14 ± 0.04, *p* = 0.005) compared to each reference group. Detailed reports of all regression analyses are shown in Supplementary Table 7.

**Figure 4.**
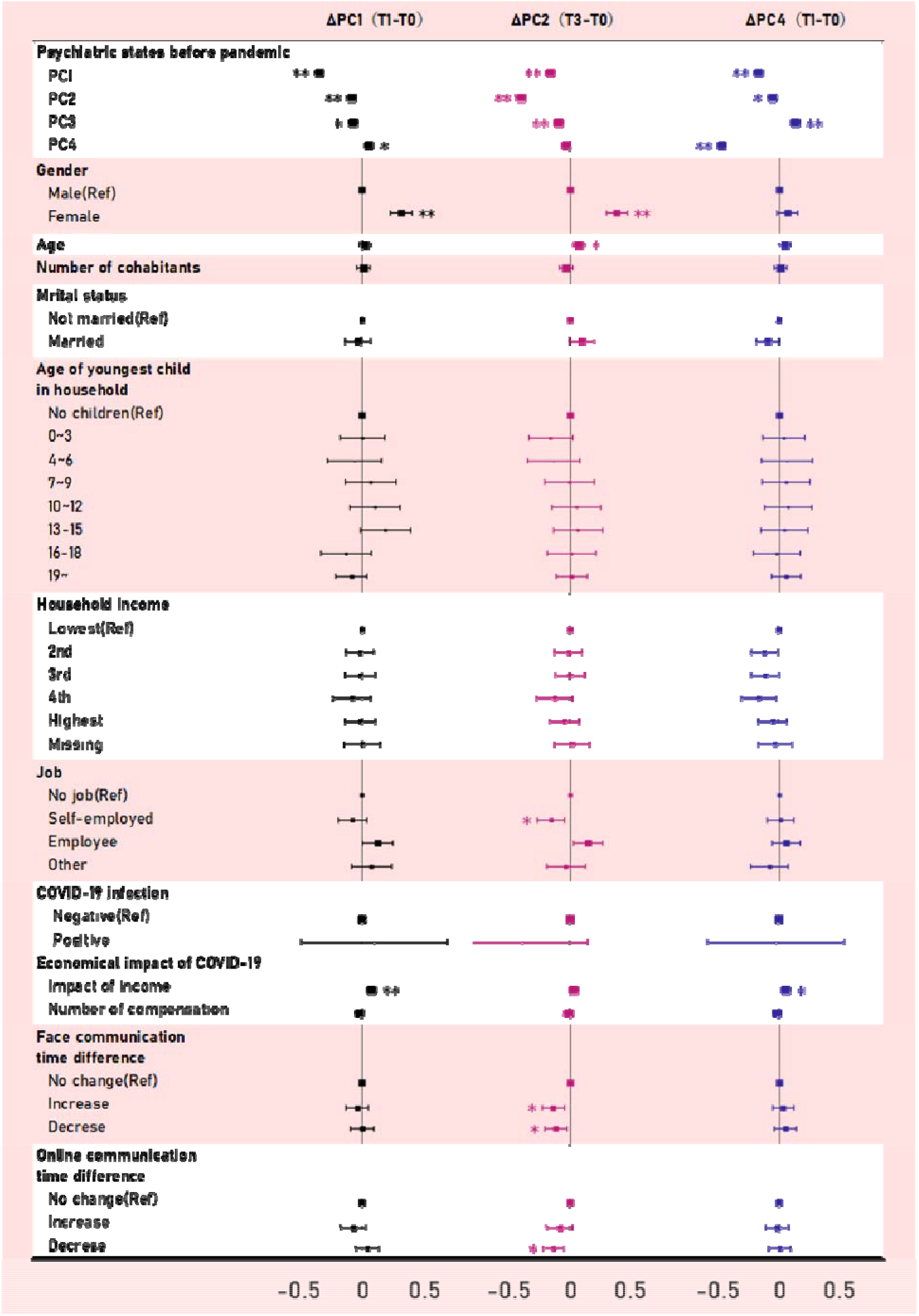
Fixed-effects regression analyses showing within-person changes in multi-dimensional psychiatric status during the pandemic.

To assess the influence of participants who dropped out of the study before completion, all analyses were also performed in the “survey population”, which included individuals with missing data (Figure 1). Results are consistent with analyses of participants with no missing data, except for the effect of age in PC4 (Supplementary Figure 4 and Supplementary Table 8).

## DISCUSSION

This is the first study to examine multidimensional psychiatric states at multiple time points from before the COVID-19 pandemic to one year after the outbreak (T0: December 2019, T1: August 2020, T2: December 2020, T3: April 2021) in a large online population (including participants with missing values: N = 3935, excluding those with missing values: N = 2274). Average psychiatric disorder scores showed different trajectories across disorders while being correlated within participants. As a result, these psychiatric dimensions were aggregated into four major, orthogonal principal components (PCs). PC1, PC2, and PC4 showed distinct exacerbation trajectories, as well as distinct peaks. PC3 showed no significant change during the pandemic.

The PCs are interpreted as general psychiatric burden, social withdrawal, alcohol-related problems, and depression/anxiety. Most psychiatric disorders contributed to PC1, which may reflect the general psychiatric burden due to the pandemic. PC2 was mainly associated with fear and avoidance aspects of social anxiety, as well as internet-related problems, which may represent symptomatic social withdrawal. Exacerbation of PC2 may result from strategies to prevent spreading of the infection, such as social distancing. Repeated avoidance of social activities may gradually instill the notion that social communication is immoral or something to be avoided. Severe social withdrawal—*hikikomori*—has been an increasing social problem in Japan. Hikikomori is characterized by a tendency to isolate oneself from society, to stay in one’s room, and to become dependent on the internet and games [42, 43]. Internet-related problems and social anxiety are gaining attention as important risk factors for social withdrawal [44–46]. PC4 was mainly associated with depression and anxiety. These PCs peaked at different stages of the pandemic. PC1 and PC4 peaked in the first stage of the pandemic. PC2 peaked with a delay, during the pandemic.

In our analysis, each psychiatric disorder was decomposed into different PCs that evolved along different trajectories. For example, alcohol-related problems contribute significantly to both PC3 and PC4. PC4, a component mainly reflecting depression/anxiety, worsened during the pandemic. PC3, a component mainly reflecting alcohol-related problems, did not change significantly during the pandemic. These data suggest that while alcohol-related problems did not display pandemic-induced changes, some individuals used alcohol maladaptively to cope with depression/anxiety. Thus, when assessing changes in mental illness due to effects of a pandemic, it is important to consider multiple dimensions to identify risks, rather than assessing single dimensions.

Our regression analysis further highlighted the importance of flexible countermeasures for mental health problems. Some risk factors were shared across PCs. Specifically, being female was a common risk factor for exacerbation of psychiatric disorders represented by the PCs. Japanese females also experienced a severe increase in suicide during the pandemic in comparison with Japanese males [47]. There is an urgent need for counter-measures to reduce the physical and mental burdens imposed by the pandemic. In parallel, we identified risks and protective factors that were specific for each PC. General psychiatric burden and depression/anxiety, PC1 and PC4, were strongly influenced by economic factors, whereas social withdrawal, PC2, was strongly influenced by human relationships. The effects of reduced income on mental health are consistent with a previous study reporting individual economic damage as a risk factor for worsening mental well-being during the pandemic [15].

In the social withdrawal component, self-employment and changes in communication were protective factors. Employment in isolation has been associated with higher social isolation during COVID-19 [48], but self-employment may be associated with lower social isolation due to COVID-19. To preserve mental well-being, a successful strategy might involve focusing on countermeasures against economic impact in the early stages of the pandemic, while supporting social interaction may be more important in later stages of the pandemic.

Given the complex nature of the link between the current pandemic and mental health, this study has some limitations. First, long-term effects of this pandemic cannot be assessed yet. Second, the psychiatric scores assessed here did not include some important dimensions, such as psychotic symptoms. Third, our analysis focused on the Japanese population. Our understanding of the effects of the COVID-19 pandemic on mental health would benefit from international comparisons, including race, culture, religion, and psychiatric states that were not assessed in this study. Finally, because our data were concatenated across time-points, the PCs reflected both temporal covariance and covariance between participants. Future work with more time points may help us clearly distinguish these two effects, to better understand the impact of the pandemic on mental health.

In summary, time courses of nine psychiatric symptoms during the COVID-19 pandemic were aggregated into three exacerbated, orthogonal principal components with different peaks, as well as different modulating factors. Our findings underline the importance of flexible approaches for mental health protections. Long-term monitoring and real-time reporting are both necessary to determine the full consequences of COVID-19 on mental health.

## Data Availability

The main summary statistics that support the findings of this study are available in the Supplementary Data. Owing to company cohort data sharing restrictions, individual-level data cannot be publicly posted. However, data are available from the authors upon request and with permission of KDDI Corporation.

## Corresponding Authors

Toshinori Chiba, MD, Department of Decoded Neurofeedback, Computational Neuroscience Laboratories, Advanced Telecommunications Research Institute International, 2-2-2 Hikaridai, Seika-cho, Soraku-gun, Kyoto 619-0288 JAPAN.

## Author Contributions

Author Contributions: TC made substantial contributions to the study conception and design. TO, NK, YM, MH, TH, and TC made substantial contributions to data acquisition. TO, FN, TK, and TC conducted statistical analyses. TO, FN, TK and TC made substantial contributions to interpretation of data. TO, TK, FN, HT, SB, TK, MN, AC, MT, MK, and TC wrote the manuscript. All authors contributed to editing and commenting on the final version. TC takes responsibility for the integrity of the work.

## Acknowledgements

This research was supported by a KDDI collaborative research contract. It was also supported by Innovative Science and Technology Initiative for Security Grant Number JPJ004596, ATLA, Japan. We thank Rumi Yorizawa, Misa Murakami, Anna Shimafuji, and Miho Nagata for data collection and organization.

