## Supplementary information for "Multiple-time measurements of multidimensional psychiatric states from immediately before the COVID-19 pandemic to one year later: A longitudinal online survey of the Japanese population"

**Supplementary method**

**The details of the survey company**

We conducted an online survey through Macromill, Inc., the largest online survey company in Japan. This online platform has a participant pool of about 1.2 million individuals living in Japan.

**Participants exclusion criteria**

We excluded 326 individuals because of inconsistencies or contradictions in their answers as follows: sleep time is the same as wake-up time in any of the surveys; gender inconsistencies (e.g., T0 gender ≠ T1 gender); the contradiction of alcohol drink in Alcohol Use Disorders Identification Test (AUDIT) (e.g., A participant answered “Never” in 1st question “How often do you have a drink containing alcohol?”, but they also answered “3 or 4” or more in 2nd question “How many drinks containing alcohol do you have on a typical day when you are drinking?”. An additional 419 individuals were excluded because they responded identically to all items, using only the maximum or minimum values in the questionnaires, including reverse questions (i.e., in CES-D, STAI-Y, AQ).

**The details of the questionnaires**

Major depression disorder was measured using the Center for Epidemiologic Studies Depression Scale (CES-D), internal consistency of 0.85 in the original version (1). Construct validity has been confirmed by Hamilton Clinician’s Rating scale (r = 0.44) and the Raskin Rating scale(r = 0.54)(1). The reliability and validity have been confirmed in Japanese (Cronbach’s α = 0.80) (2). General anxiety was measured by the State-Trait anxiety scale (STAI)(3), which has a Cronbach’s alpha of 0.86 to 0.95 in the original study. The reliability and validity have been confirmed in Japanese (Cronbach’s α = 0.92)(4). Social anxiety was measured using Liebowitz Social Anxiety Scale (LSAS-fear/avoid) (5), a Cronbach’s alpha of 0.95 in the original version. Construct validity has been confirmed by Safren’s four-factor model in the original version (5) and Social Avoidance and Distress Scale and Professional Diagnosis in the Japanese version(6). Obsessive-compulsive disorder was measured using the Obsessive-Compulsive Inventory (OCI), a Cronbach’s alpha of 0.86 to 0.95 in the original version (7) and 0.96 in the Japanese version (8). Construct validity has been confirmed by Yale-Brown Obsessive Compulsive Scale, Compulsive Activity Checklist, and Maudslay Obsessive-Compulsive Inventory in the original version and Maudsley Obsessive-Compulsive Inventory (r = 0.74), STAI (r = 0.41), and CES-D (*r* = 0.48) in the Japanese version (8). Alcohol dependence was measured using the Alcohol Use Disorders Identification Test (AUDIT), a Cronbach’s alpha of 0.65-0.93 in the original version (9). The reliability and validity have been confirmed in Japanese (Cronbach’s α = 0.81, construct validity was confirmed by structure interview (10). Internet-related problems were measured using the Compulsive Internet Use Scale (CIUS), a Cronbach’s alpha of 0.89 (11) in the original version and more than 0.9 in the Japanese version (12). Construct validity has been confirmed by the strong positive correlation with the Online Cognition Scale (*r* = 0.70, *p* < 0.001) and the amount of time spent online (r = 0.33, *p* < 0.001) in the original version and POSI, MR, DSR, NO, K6 and UCLA Loneliness in the Japanese version. Autism Spectrum Disorder (ASD) was measured using the Autism-Spectrum Quotient (AQ), a Cronbach’s alpha of 0.63~0.77 in the original version(13) and internal consistency of 0.81 in the Japanese version (14). Construct validity has been confirmed by the professional diagnosis based on DSM-Ⅳ (*r* = 0.58) and the experience of exposure to social adaptation problems (*r* = 0.92) in the Japanese version (14). Adult attention-deficit/hyperactivity disorder (ADHD) was measured using the Adult ADHD Self-Report Scale(ASRS) (15), a Cronbach’s alpha of 0.88 to 0.89 in the original version (16), 0.83 in the Japanese version (17). Construct validity has been confirmed by the Japanese version of the Conners’ Adult ADHD Rating Scales-Self Report and Beck Depression Inventory-Ⅱ in the Japanese version (17).

For the original study, we assessed other scales and excluded them from the analysis to reduce the redundancies in the variables. STAI-Y-trait, Internet gaming disorder (IGDS), and Internet addiction test (IAT) were also assessed for the original study purpose. In this study, these scores were excluded from the analysis to reduce the redundancies in the variables: STAI-Y-trait was excluded because we measured the STAI-Y-state. IGDS-J and IAT were excluded because we measured the CIUS.

**Supplementary Table 1** A list of the COVID-19 pandemic related questions

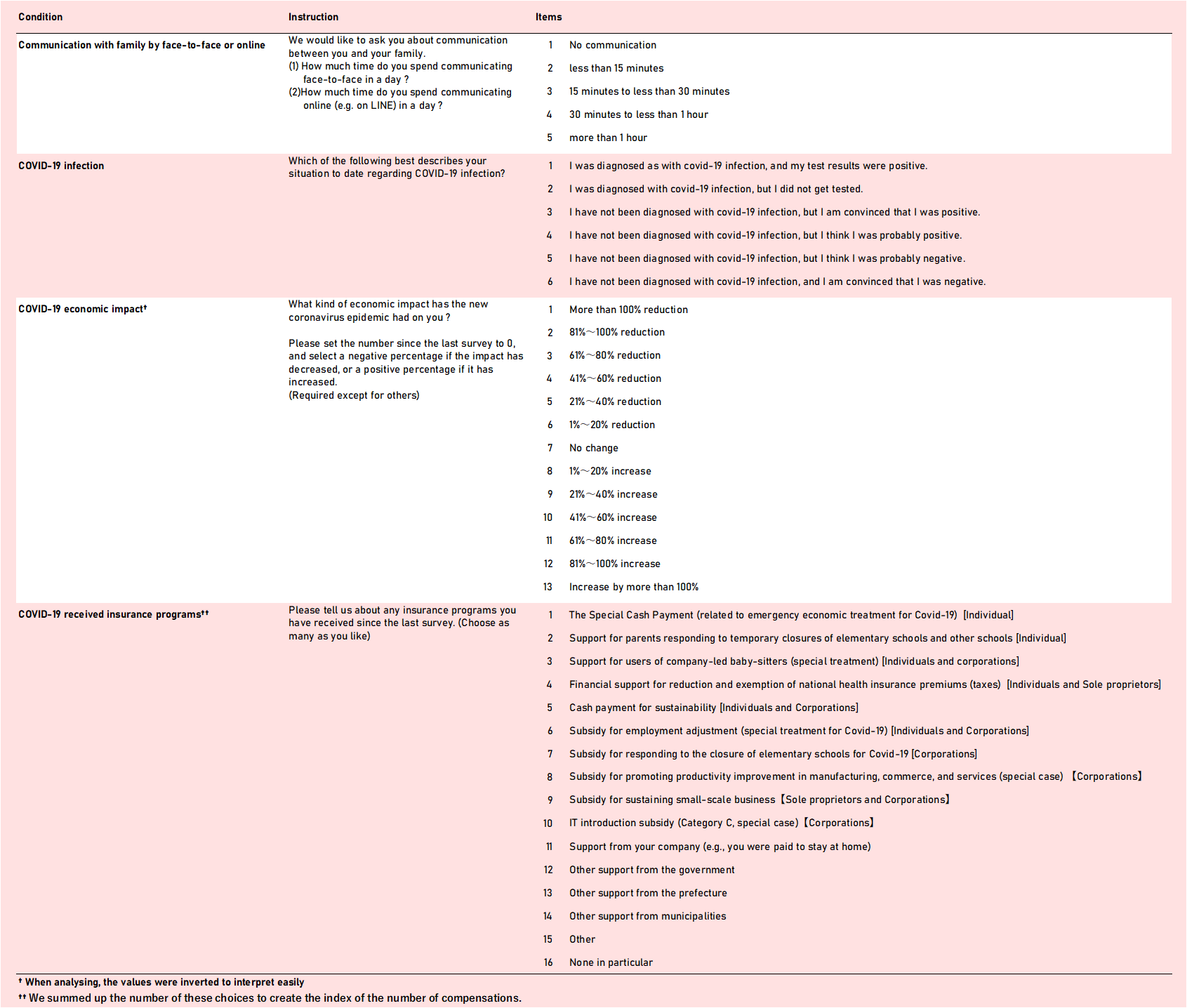

**Supplementary Table 2** Characteristics of the survey population (N = 3935)

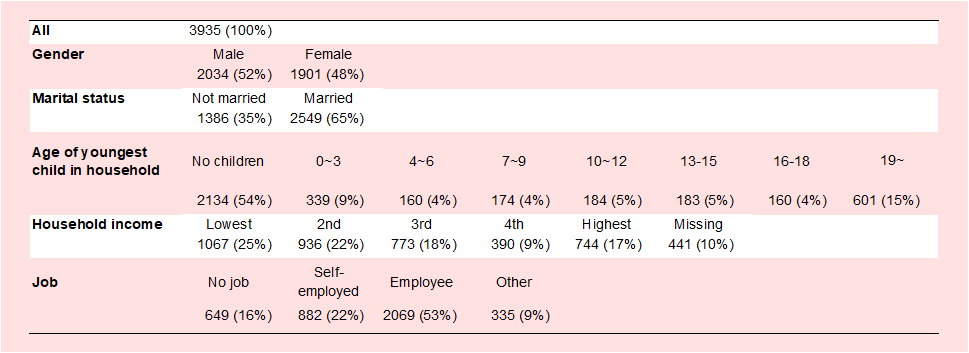

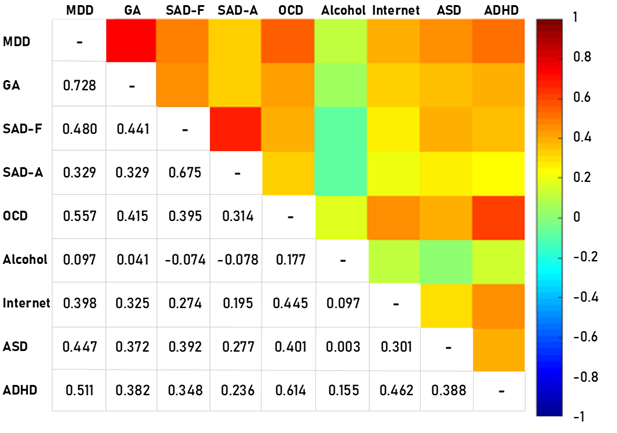

**Supplementary Figure 2** Correlation matrices for changes in scores of psychiatric disorders over time

MDD: major depressive disorder as measured by CES-D; GA: general anxiety as measured by STAI-Y; SAD-F: fear aspects of social anxiety disorder as measured by LSAS-F; SAD-A: avoidance aspects of social anxiety disorder as measured by LSAS-A; OCD: obsessive-compulsive disorder as measured by OCI; Alcohol: alcohol-related problems as measured by AUDIT; Internet: internet-related problems as measured by CIUS; ASD: autism as measured by AQ; ADHD: attention deficit and hyperactivity disorders as measured by ASRS.

**Supplementary Table 3** *p* values of each psychiatric exacerbation

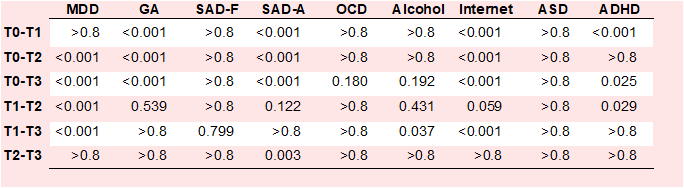

MDD: major depressive disorder as measured by CES-D; GA: general anxiety as measured by STAI-Y; SAD-F: fear aspects of social anxiety disorder as measured by LSAS-F; SAD-A: avoidance aspects of social anxiety disorder as measured by LSAS-A; OCD: obsessive-compulsive disorder as measured by OCI; Alcohol: alcohol-related problems as measured by AUDIT; Internet: internet-related problems as measured by CIUS; ASD: autism as measured by AQ; ADHD: attention deficit and hyperactivity disorders as measured by ASRS. Tx-Tx represents the difference between two-time points. *P* values were adjusted by Bonferroni correction.

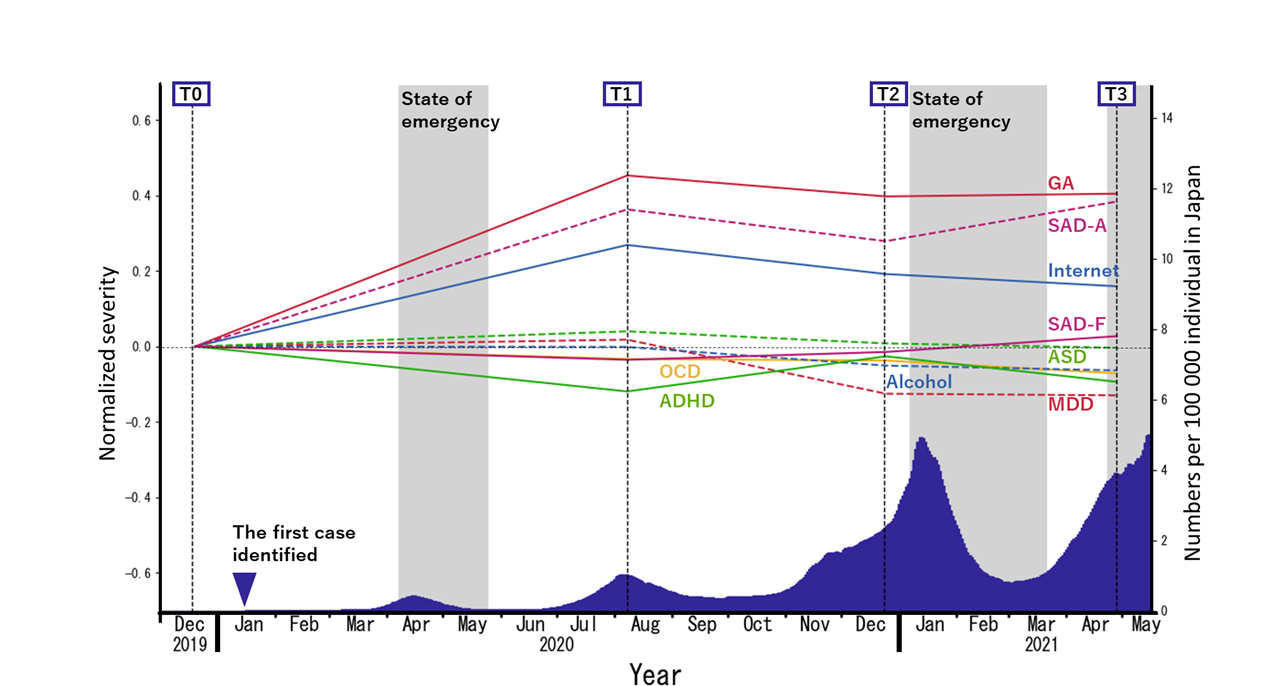

**Supplementary Figure 2** The trajectories of each psychiatric score in the survey population (N = 3935)

The blue area under the curve represents a 15-day moving average of daily new cases of COVID-19 per 100 000 Japanese residents.

GA: general anxiety as measured by STAI-Y; SAD-A: avoidance aspects of social anxiety disorder as measured by LSAS-A; Internet: internet-related problems as measured by CIUS; ASD: autism as measured by AQ; SAD-F: fear aspects of social anxiety disorder as measured by LSAS-F; OCD: obsessive-compulsive disorder as measured by OCI; ADHD: attention deficit and hyperactivity disorders as measured by ASRS; Alcohol: alcohol-related problems as measured by AUDIT; MDD: major depressive disorder as measured by CES-D.

**Supplementary Table 4** Characteristics of the study population and each PC loading in the study population (N = 2274)

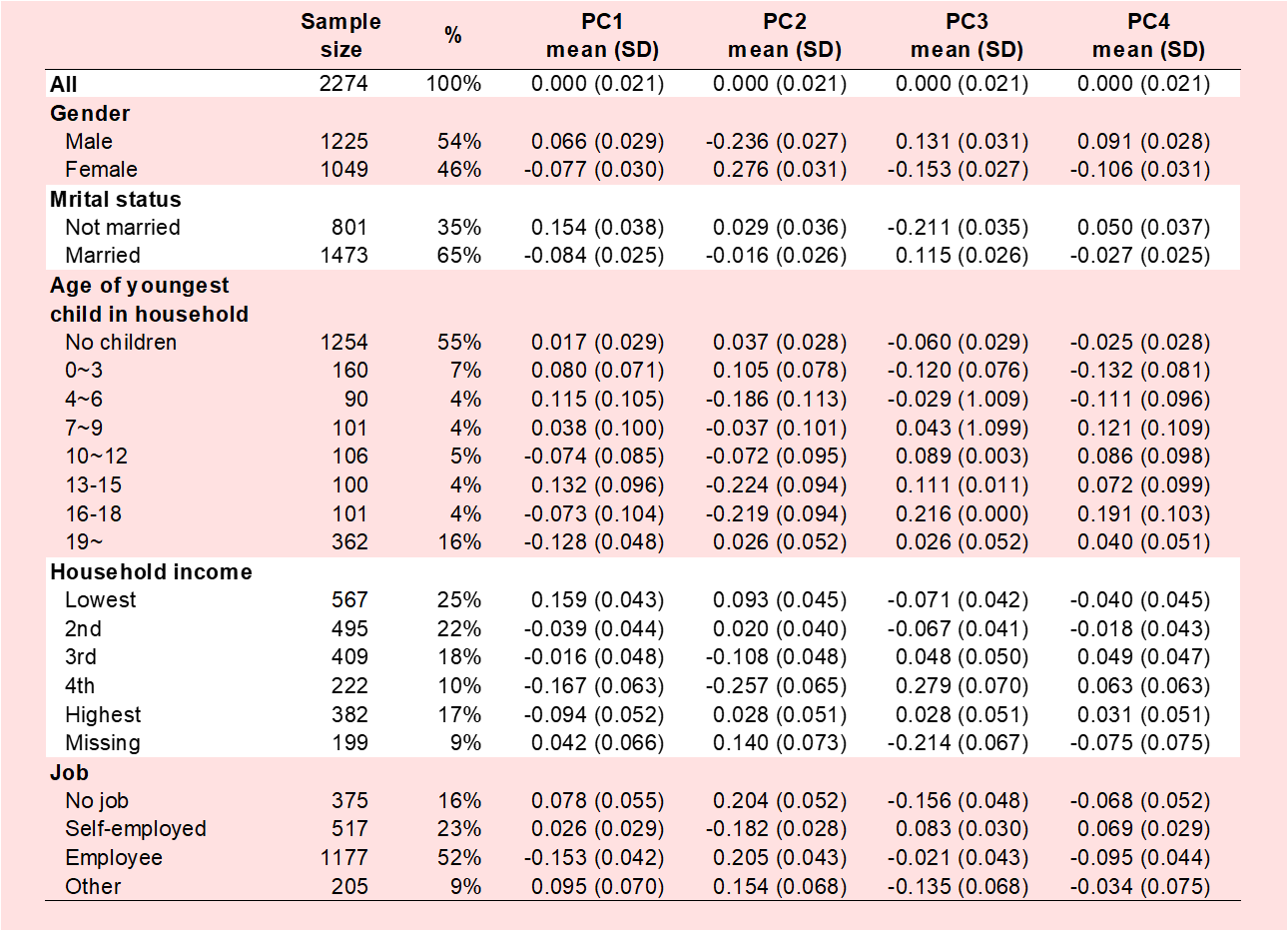

PC1: Principal component 1; PC2: Principal component 2; PC3: Principal component 3; PC4: Principal component 4.

**Supplementary Table 5** Characteristics of the study population and each PC loading in the survey population (N = 3935)

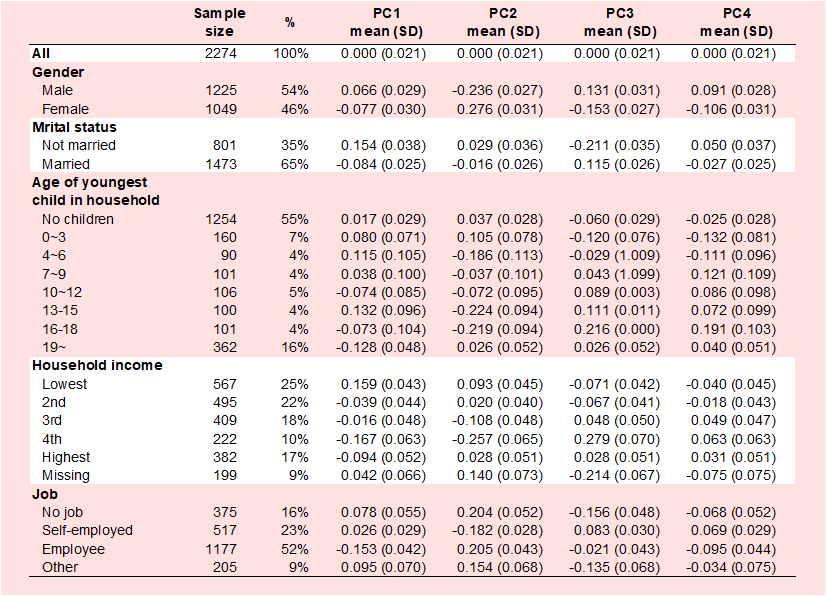

PC1: Principal component 1; PC2: Principal component 2; PC3: Principal component 3; PC4: Principal component 4.

**Supplementary table 6** *p* values of each principal component during the pandemic

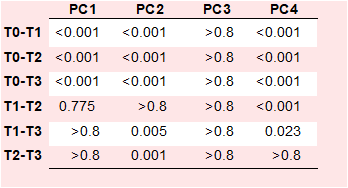

PC1: Principal component 1; PC2: Principal component 2; PC3: Principal component 3; PC4: Principal component 4. *p* values were adjusted by Bonferroni correction.

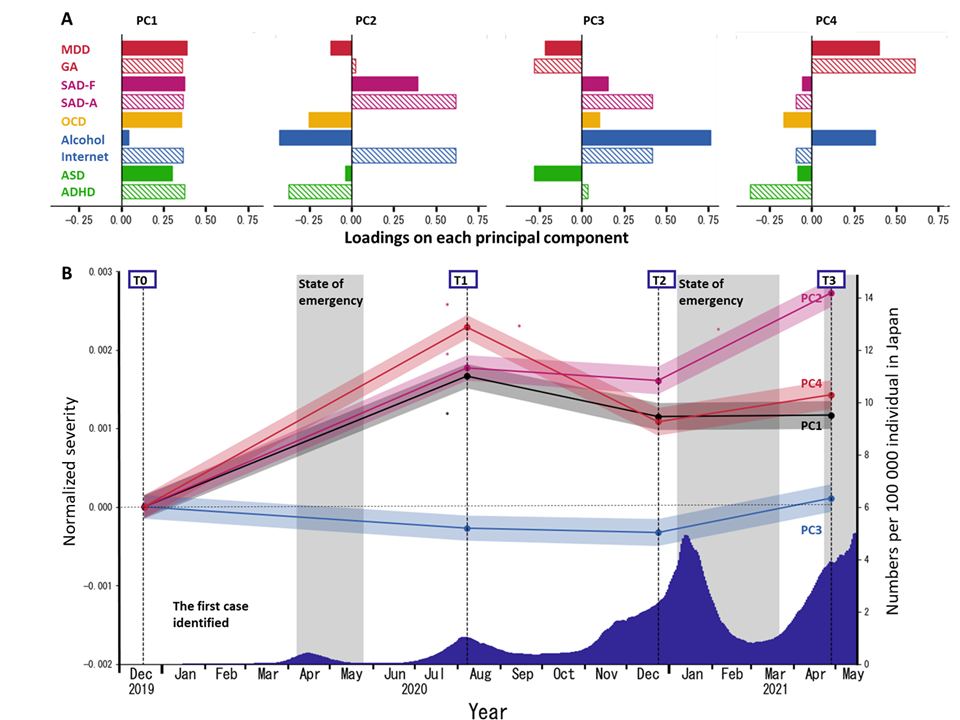

**Supplementary Figure 3** The trajectories of PC scores obtained from multidimensional psychiatric scores in the survey population (N = 3935)
**A.** Loadings of psychiatric disorder scores for each principal component. **B.** Trajectories of the average of each PC is shown. The blue area under the curve represents a 15-day moving average of daily new COVID-19 cases per 100 000 Japanese residents. Signs of PCs were arranged so that each maximum loading assumed a positive value. Asterisks indicate significant changes in PC score from the previous time point (p<.05, Bonferroni-corrected). All PC scores during the pandemic (T1, T2, and T3) except those of PC3 are significantly higher than the scores pre-pandemic (T0).

MDD: major depressive disorder as measured by CES-D; GA: general anxiety as measured by STAI-Y state, SAD-F: fear aspects of social anxiety disorder as measured by LSAS-F; SAD-A: avoidance aspects of social anxiety disorder as measured by LSAS-A; OCD: obsessive-compulsive disorder as measured by OCI; Alcohol: alcohol-related problems as measured by AUDIT; Internet: internet-related problems as measured by CIUS; ASD: autism as measured by AQ; ADHD: attention deficit and hyperactivity disorders as measured by ASRS

**Supplementary Table 7** The statistical values of fixed-effects regression analyses showing the within-person changes in multi-dimensional psychiatric status with the pandemic in the study population (N = 2274).

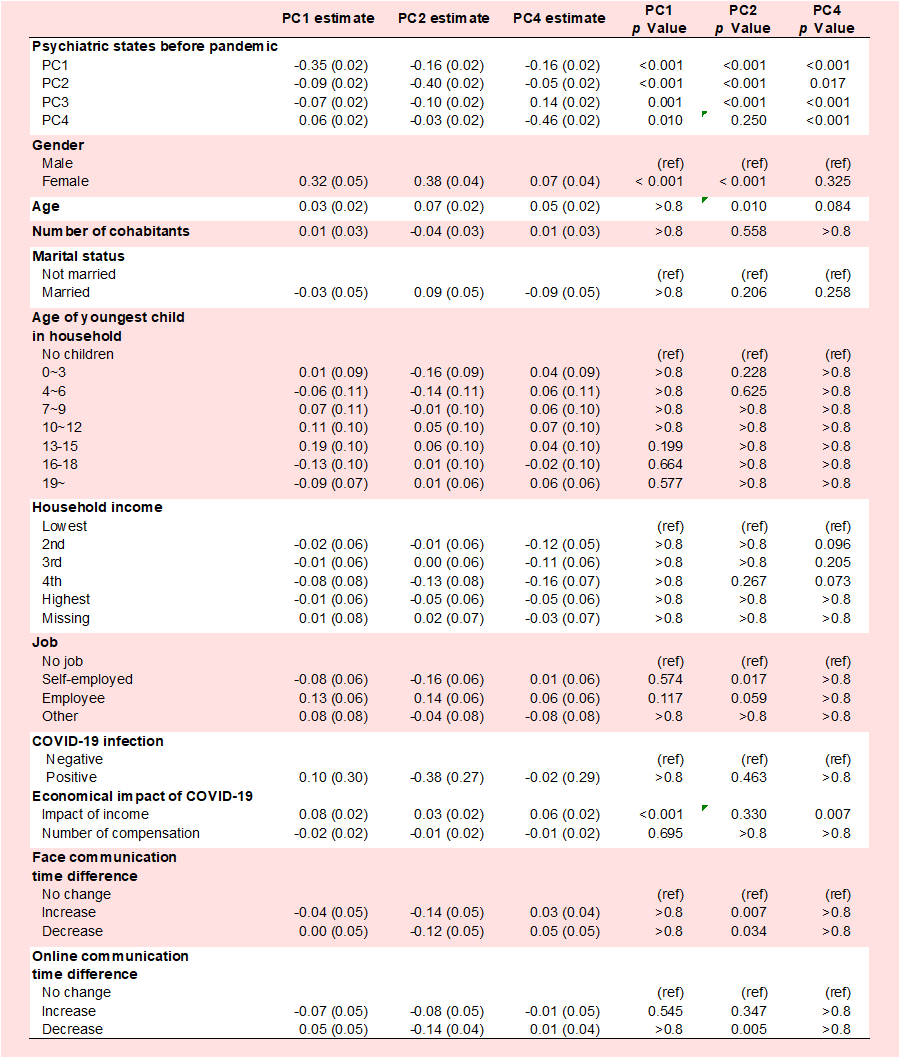

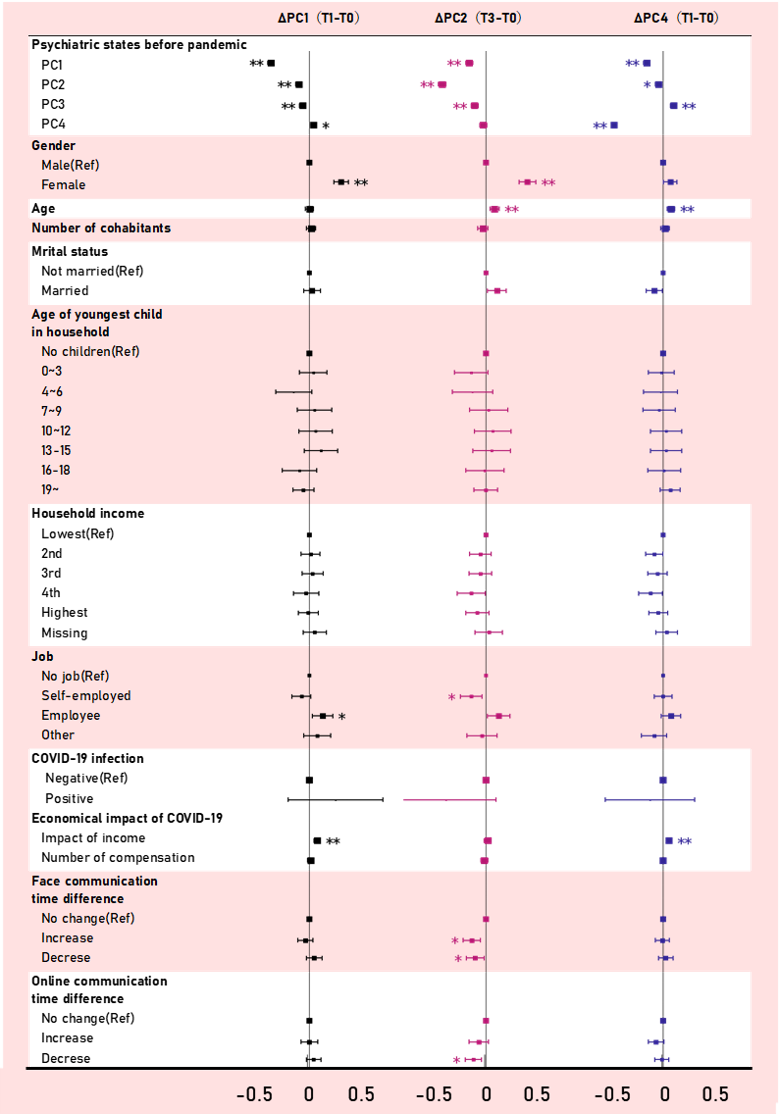

**Supplementary Figure 4** The fixed-effects regression analyses showing the within-person changes in multi-dimensional psychiatric status with the pandemic in the survey population (N = 3935)

**Supplementary Table 8** The statistical values of fixed-effects regression analyses showing the within-person changes in multi-dimensional psychiatric status with the pandemic in the survey population (N = 3935).

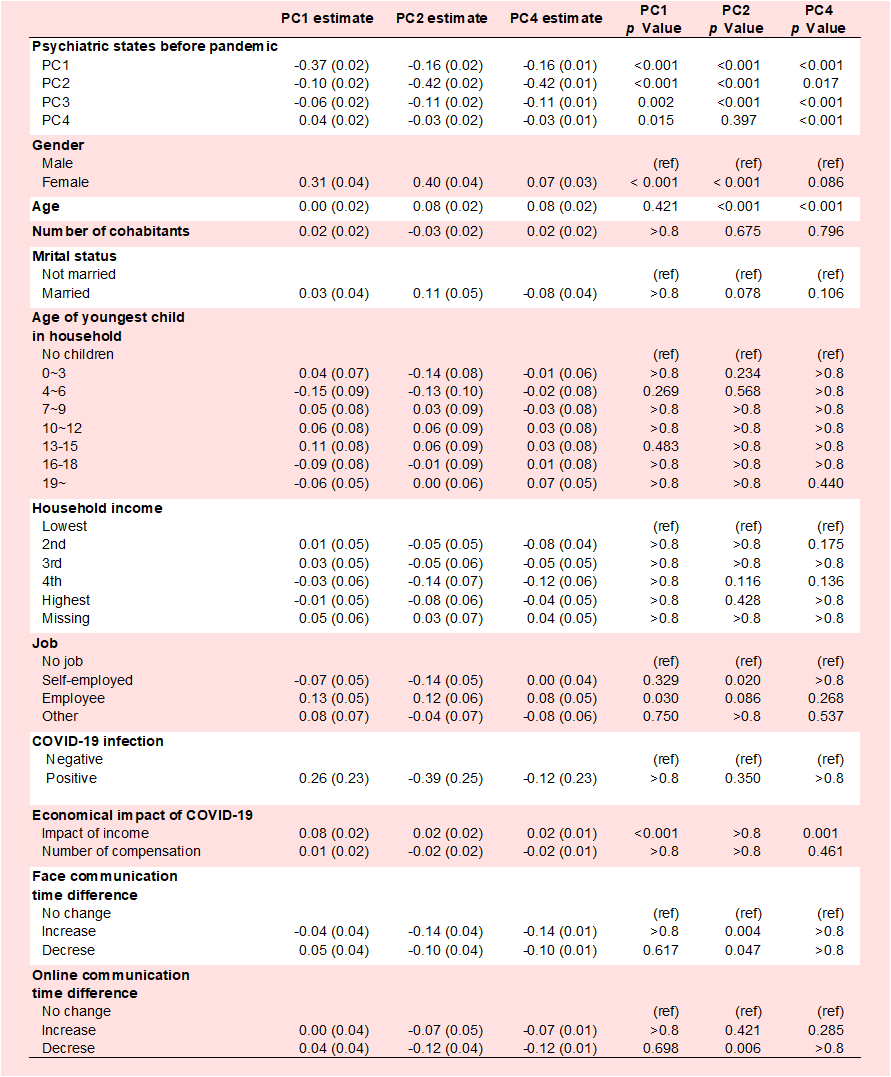
